# Academic and socio-professional trajectories in narcolepsy type 1: the NARCOSCOL-NARCOVITAE multicentric study

**DOI:** 10.1101/2024.01.13.24301265

**Authors:** Laure Peter-Derex, Emmanuel Fort, Benjamin Putois, Nora Martel, François Ricordeau, Hélène Bastuji, Isabelle Arnulf, Lucie Barateau, Patrice Bourgin, Yves Dauvilliers, Rachel Debs, Pauline Dodet, Benjamin Dudoignon, Patricia Franco, Sarah Hartley, Isabelle Lambert, Michel Lecendreux, Antoine Léotard, Smaranda Leu-Semenescu, Laurene Leclair-Visonneau, Damien Léger, Martine Lemesle-Martin, Nadège Limousin, Régis Lopez, Nicole Meslier, Jean-Arthur Micoulaud-Franchi, Christelle Charley-Mocana, Marie-Pia d’Ortho, Pierre Philip, Elisabeth Ruppert, Sylvie de La Tullaye, Manon Brigandet, Jennnifer Margier, Benjamin Rolland, Barbara Charbotel, Stéphanie Mazza

## Abstract

**Background and objectives:** Narcolepsy type 1 (NT1) is a chronic, disabling neurological disease. Sleep-related symptoms and comorbidities such as psycho-cognitive disturbances, and a frequent childhood onset of the disease may negatively impact patients’ career. We conducted a multicentric comparative cross-sectional study in Reference/Competence Centers for Narcolepsy in France to investigate the educational and occupational paths of patients with NT1.

**Methods:** Between February 2020 and 2023, adult patients with NT1 regularly followed-up in the participating centers were invited to complete online questionnaires including the Epworth sleepiness Scale, Narcolepsy Severity Scale, Beck Depression Inventory II, Siegrist questionnaire, Adult Self-Report and Adult Behavior Checklist, and a customized questionnaire on academic and professional trajectories. Controls were selected from within the patients’ close circle. Comparisons were adjusted for sex and age, and the determinants of patients’ professional prognosis were quantified by a composite score including professional-related outcomes.

**Results:** Questionnaires were filled by 235 patients (63.8% women, 36.4±14.7 years, 86.5% treated, 66.4% with childhood onset) and 166 controls (69.9% women, 40.3±14.4 years). No difference was observed between patients and controls for grade repetition and graduation level distribution, but patients reported more interruptions in their scolarity which was considered difficult, with more absenteeism and lateness. No difference was observed for employment rate (69.5% vs 77.0%) and socio-professional category distribution, but income was lower in patients who reported more unwanted changes in position and part-time work, with increased effort-reward imbalance (OR=2.28 95% CI [1.20-4.33], p=0.01). Almost half of the patients benefited from an official disability recognition and 10.2% received invalidity benefits. Impaired professional prognosis was associated with depression (p<0.0001) and attention disorders (p=0.03), while being narcoleptic during schooling was a protective factor (p=0.02). Residual sleep-related symptoms were not significant predictors.

**Discussion:** Most patients with NT1 manage to achieve their careers goals, but at the cost of an effort/reward imbalance. Early diagnosis during childhood might allow a better adjustment to the disease. The critical role of co-morbidities in professional trajectories suggests that, in treated patients, psycho-cognitive disturbances have greater impact on daily functionning than sleep-related symptoms, and stresses the need to consider psycho-cognitivo-social dimensions in patient care.

## Introduction

Narcolepsy type 1 (NT1) is a rare (prevalence 0.025 to 0.05%) neurological disease caused by a deficiency in orexin, a hypothalamic neurotransmitter involved in sleep/wake regulation.^1, 2^ Patients with NT1 present with hypersomnolence, disrupted nighttime sleep, and symptoms related to rapid-eye-movement (REM) sleep dysregulation such as cataplexy (transient loss of muscle tone triggered by emotions), sleep paralysis, and sleep-related hallucinations.^2^ Importantly, several comorbidities impair daytime functioning and quality of life in NT1 patients,^3, 4^ including cognitive impairments (mainly attentional deficit but also subtle alterations in other domains such as decision-making and emotional processing),^5^ anxiety/depression,^6^ early-onset puberty, and obesity as well as altered metabolic profile.^7^ These multiple symptoms, the frequent onset of the disease in childhood or young adulthood and the fact that narcolepsy is a chronic condition explain why school and work issues are major challenges in the care of patients with NT1.

Regarding academic progress, several works have highlighted the burden of narcolepsy at school despite normal or high intelligence quotient scores.^8–10^ Poor academic performance, increase in grade repetition, and absenteeism were reported in most studies in children and adolescents.^11, 12^ Sleepiness may contribute to educational difficulties^13^ although behavioral, emotional and cognitive aspects may also be involved.^8, 14, 15^ Interestingly, academic performance has been found to improve after appropriate treatment.^16, 17^ Data retrieved from adult patients are in line with these pediatric studies, even if some research provides a more nuanced perspective by suggesting that, despite academic difficulties, the graduation rate might not be significantly impaired. Three studies reported no difference in educational level in adults with narcolepsy as compared to healthy controls or the general population,^14, 18, 19^ whereas significant impairment was reported in patients with childhood and adolescent-onset narcolepsy.^20^ Regarding professional progress, trajectories seem more difficult for patients, although available data are also heterogeneous. Employment rates lower than those of control subjects were reported in some studies (41%,^21^ 47%,^22^ 49%^4^), while others found no significant impairment (57%,^19^ 59%,^23^ 67%,^18^ 73%,^24^ 82%^14^). Such heterogeneity between studies may reflect methodological differences in several domains: data collection (registers vs questionnaires vs direct interviews); characteristics of control and patient populations, especially regarding the proportion of NT1 vs narcolepsy type 2 (NT2), the ratio of childhood and adulthood disease onset, and the age at the time of the study, which appear to be critical factors^2, 11, 18^; and the period during which the studies were carried out, with a possible improvement in diagnosis and treatment over the last decades.^25^ Nevertheless, beyond quantitative global outcomes, most studies agree that there are qualitative alterations of dimensions linked to educational and occupational areas. In children, several works have highlighted interpersonal problems with teachers and peers, feelings of being misunderstood, bullying and stigmatization.^9, 14^ In adults, reduction in work productivity, increased rates of absenteeism and presenteeism, fewer promotions, and fear of job loss are almost constantly reported.^4, 14, 20–22, 24, 26, 27^ This issue is all the more important as employment status and career advancement strongly influence quality of life of patients with narcolepsy.^23, 28, 29^ Last, studies are unanimous on the economic impact of the disease, in terms of both direct and indirect costs, and in particular the reduction in patients’ income.^4, 20–22, 26^

Recently, we reported the results of the Narcowork single-center study, which included adult patients with NT1 and NT2.^19^ Although we found no difference between patients and controls with respect to grade repetition, highest graduation, socio-professional category (SPC) and employment status, both academic and occupational paths were experienced as difficult by patients. In addition, worse professional prognosis was observed for NT1 compared to NT2 patients and controls. This pilot study prompted us to conduct a larger multicentric study in France, the NarcoScol-NarcoVitae study, with a similar methodology but focused on NT1 patients, in order to refine these findings. We aimed to (i) compare academic and professional outcomes in NT1 patients vs healthy controls (ii) describe adaptations at school and in the workplace (iii) identify prognostic factors linked to professional outcomes.

## Methods

### Study design and participants

The NarcoScol-NarcoVitae multicentric comparative cross-sectional study was conducted in the Reference and Competence Centers for Narcolepsy and Rare Hypersomnia Network in France (n=18 centers) between February 2020 and February 2023. Adult patients followed up in participating centers and diagnosed with NT1 according to the International Classification of Sleep Disorders, third Ed. (ICSD-3) criteria (excessive daytime time sleepiness over 3 months + cataplexy and a positive multiple sleep latency test [sleep latency ≤8 minutes and >1 REM sleep period within 15 min of sleep onset] OR low orexin-A cerebrospinal fluid levels [< 110pg/mL]) were eligible.^30^ Exclusion criteria were: insufficient knowledge of the French language, cognitive impairment inconsistent with questionnaire completion and refusal to participate. Patients were informed of the study via letter by the investigator of the Center where they were usually followed up, and invited to fill out online questionnaires on a dedicated website. They were asked to find two acquaintances (not relatives, same sex and age [±5 years]) willing to complete the same questionnaires except for those specific to narcolepsy. The aim of this procedure was to find controls socio-demographically matched to the patients, and motivated to complete the questionnaires. As part of routine follow-up in Narcolepsy centers, all patients were approached by the investigator at each consultation to re-inform them about the study. On this occasion, a minimum dataset was directly collected in a registry, including age, sex, highest graduation and current professional status.

### Questionnaires

The questionnaires included:

- Validated questionnaires: Epworth sleepiness Scale (ESS)^31^, Beck Depression Inventory II (BDI II)^32^, the Narcolepsy Severity Scale (NSS)^33^, Siegrist questionnaire to assess the effort-reward imbalance at work^34^, EuroQol quality of life scale (EQ5D-5L)^35^, and the Adult Self-Report (ASR) and Adult Behavior Checklist (ABCL, completed by a relative or close friend)^36^ designed to evaluate behavioral, psychosocial and adaptive functioning in adults.
- A customized questionnaire including (details in supplementary Table 1): demographic information; medical history and treatments; use of alcohol, tobacco, psychostimulant substances, and cannabis; narcolepsy characteristics; physical activity; leisure/family/friends activities; academic and professional trajectories, disability status. To note, in France, people with a disability can either benefit from the official recognition of disability (they work but benefit from specific adaptations at the workplace following the recommendations of the occupational physician), or, if the disability is severe, they can benefit from the “invalidity” status, i.e. a disability limiting their capacity to work, in which case they receive an insurance pension.

The time taken to complete the questionnaires was approximately 75 minutes, and patients were allowed to complete them in several sessions.

### Outcomes

The following outcomes were compared between patients with NT1 and controls: the distribution of occupational status (employed [fixed- or indefinite-term contract, self-employed], unemployed [loss of job, housewife/househusband, disability limiting work], Retired, or Student/trainee), level of graduation and grade repetition (to note, these outcomes regarding occupational status and graduation were available for patients both from the questionnaires completed by the participants and from the registry completed by the investigator), distribution of SPC, health in cluding psycho-behavioral outcomes, quality of life, academic and professional trajectories, satisfaction during academic training and at work, effort-reward imbalance at work, as well as individual and collective cost of work absenteeism and disability in the year prior to the study. Participants’ loss of income was calculated based on the number of days of work stoppage multiplied by the average net daily wage in the private sector in 2021 per SPC, from which the daily insurance pension received was subtracted. The human capital method was used to assess societal costs associated with reduced labor supply. Off-work days were valued either using 2 approaches: gross domestic product (GDP) or gross loaded salary plus employers’ social security contributions (average salary in the private sector in 2021).

In patients with NT1, we also collected details about adaptations at school and in the workplace, and feelings and experiences related to being narcoleptic at school and work.

In order to explore factors associated with a better or easier professional path, a composite score was used as previously published.^19^ This score includes the level of graduation, the cumulate duration of periods of unemployment, the number of job changes due to the disease or dismissal, the current professional status, the frequency of being late for work and the number of sick leave days during the past year; it ranges from 0 (lowest prognosis) to 6 (best prognosis).

### Statistical analysis

Descriptive statistics were presented as n (%) for categorical variables and as mean ± sd, median (Q1, Q3) for continuous variables.

Comparison between narcoleptic patients from the registry and those who had completed the questionnaires, and between cases (narcoleptic patients from the questionnaires) and controls were performed with Chi-squared test for ≥ 2 modality nominal variables or Fisher test if Chi-squared test was not applicable and with T-test test for quantitative variable or Wilcoxon when T-test was not applicable (small group size). Logistic regressions were performed in order to calculate odds ratio adjusted for sex and age. Three categories of narcoleptics patients were created, according to the age at the symptoms onset and at narcolepsy diagnostic: patients with NT1 onset and diagnosis <18 years (“group 1”), patients with NT1 onset <18 and a diagnosis ≥18 years (“group 2”) and patients with NT1 onset ≥18 (“group 3”). We used an Ordinal Logistic Regression to compare variables between these 3 categories. Multiple regressions were performed with age (quantitative) as the adjusted dependent variable. For health-economic analyses, comparison were performed with a generalized linear model and a tobit model according to the distribution of the data in order to calculate p-value adjusted for sex and age. Sensitivity analyses were performed by opting for the median and mean salary in the French population for all respondents rather than a salary based on SPC. Sub-group analyses were also conducted according to the age at disease onset and diagnosis (3 groups) and among respondents who had had at least one break during the year.

The determinants of narcoleptic patients’ professional insertion, as assessed by the composite score, were studied with a generalized linear model. A multiple stage analysis plan was conducted: first, a univariate matched analysis was performed with the following factors: age, sex, BMI, sleepiness (ESS), narcolepsy severity (NSS), depressive symptoms (BDI-II), quality of life (EQ5D-5L visual analogic scale[VAS]), ASR total score, attention disorders (ASR syndrome scale “attention issues”: 0 “clinical”, 1 “limit”, 2 “normal” ranges), effort-reward imbalance (Siegrist), number and type of treatments, age at onset and at diagnostic, diagnosis delay, patient group (according to age at disease onset and diagnosis), being narcoleptic during academic path, official recognition of disability at school/work, and specific support at school/work. Afterwards, a multivariate matched analysis was performed, which allowed to identify one entry determinant in the model of 10% and a descending variables selection.

A p value ≤ 0.05 was considered as statistically significant. Analyses were performed with SAS software (version 9.4).

### Standard Protocol Approvals, Registrations and Patient Consents

The study was performed in accordance with the principles of good clinical practice and the Declaration of Helsinki. The trial was approved by the Ethic Review Board (CPP Nord-Ouest I, N° 2018-A01586-49), and by the National Commission for Data Protection. All participants provided informed consent. The trial was registered at ClinicalTrials.gov (NCT 03765892).

### Data availability statement

The data supporting this study are available upon reasonable request to the corresponding author and in compliance with local ethical regulations.

## Results

### Participants

A total of 1,560 letters were sent to all NT1 patients registered in the active files of participating centers. Among them, 235 (63.8% women, 36.4±14.7 years) completed the questionnaires, along with 166 controls (69.9% women [p=0.2], 40.3±14.4 years [p=0.009]). The registry included 1,164 patients (53.0% women [vs patients from questionnaire: p=0.002], 38.6±15.5 years [p=0.04]), among whom data on occupational status were available for 913 individuals, and on highest level of graduation for 656. Characteristics of the disease in patients who answered the questionnaires are shown in Table 1 and supplementary Table 2. A childhood disease onset was reported in 66.4% patients, among whom 53.9% were diagnosed before age of 18 (supplementary Figure 1). Most patients (86.5%) were taking a wake promoting and/or anti-cataplectic medication.

**Table 1.**
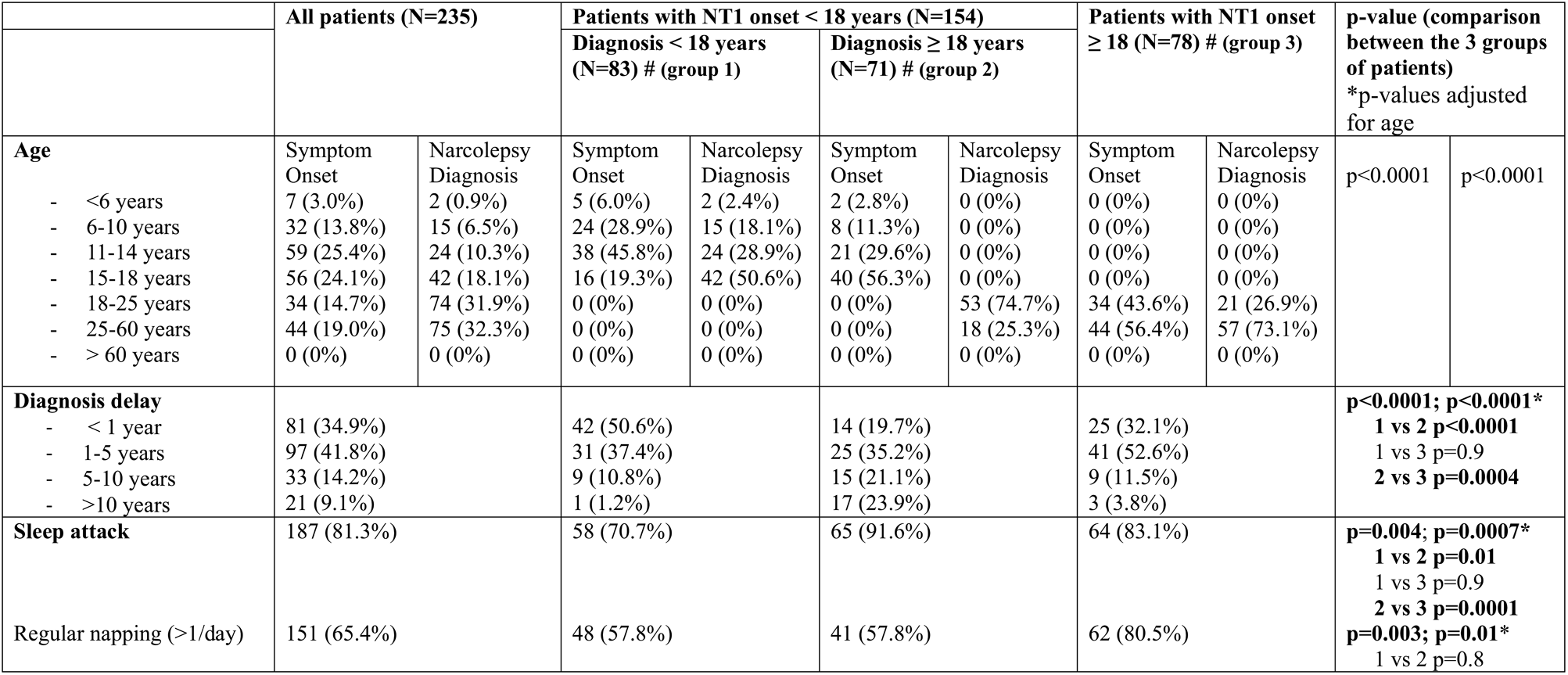
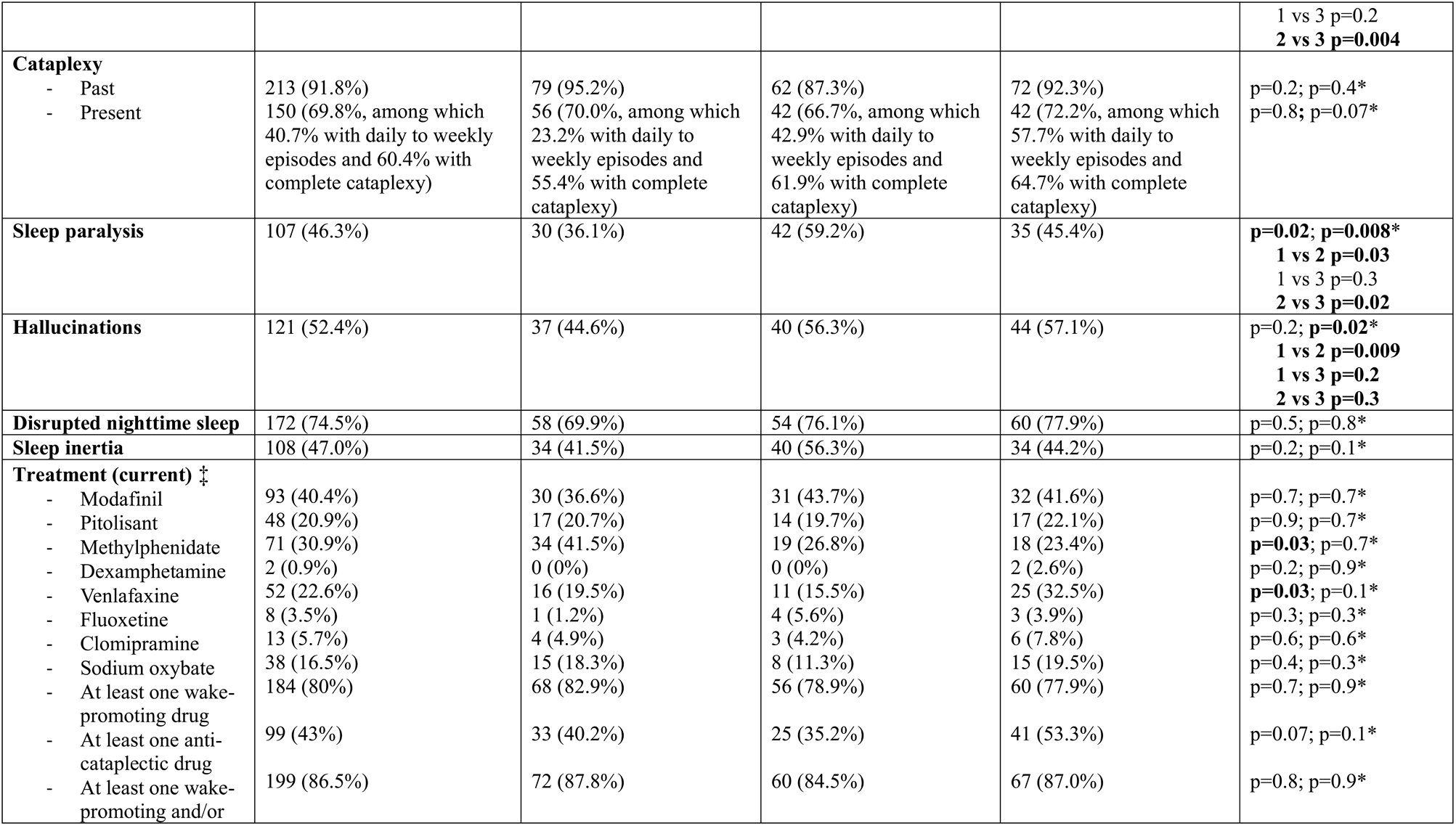
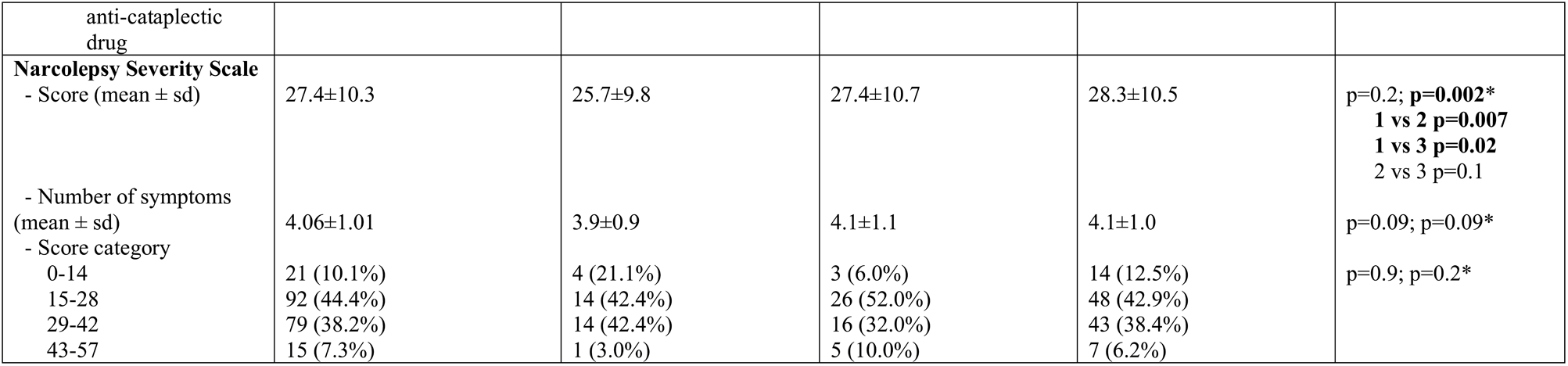
Characteristics of the disease in patients with NT1. #note that the sum of the number of patients in the 3 groups is 232, as data about age at onset and age at diagnosis were missing for 3 patients. ‡ Note that some patients were taking a combination of treatments and that some treatments (such as Solriamfetol) were not available in France at the time of the study. Comparisons between the 3 groups: significant results in bold

Socio-demographic and health-related outcomes are presented in Table 2. Patients with NT1 were less likely to be in a couple (OR=0.38 95% CI [0.22-0.66], p=0.004) and to have children (OR=0.56 95% CI [0.26-0.80], p=0.006) than controls. They also reported less physical activity (OR=0.57 CI 95% [0.38-0.88], p=0.01) and leisure time (OR=0.63 CI 95% [0.41-0.97], p=0.01). No difference between groups was found for caffeinated drink, cannabis, and alcohol consumption but the prevalence of current smoking was higher in patients (OR=1.98 95% CI [1.18-3.30], p=0.03). Regarding questionnaires, depressive symptoms were higher in patients than in controls (BDI II score 13.1±10.1 vs 8.3±6.9, p<0.0001), with a significant increase in mild (OR=1.75 95% CI [1.07-2.86]), moderate (OR=3.51 95% CI [1.64-7.49]), and severe depression (OR=23.93 95% CI [3.08-186.11]) (p<0.0001), and significantly higher subscores for pessimism, feelings of guilty and punishment, negative feelings about oneself, self-criticism, indecision, fatigue and appetite. Sleepiness was as expected higher in patients (ESS: 16.3±4.0 vs 8.3±4.9, p<0.0001), with 92.0 % of patients vs 29.4% of controls having a score >10 (OR=32.42 95% CI [17.0-63.86], p<0.0001). Quality of life was lower in patients (EQ5D-VAS 69.6±17.2% vs 81.0±13.6%, p<0.0001, OR=0.95 CI 95% [0.93-0.97], with impairment in mobility, daily life activities and anxiety/depression items. The ASR scale showed abnormal scores (“clinical range”) in patients regarding adaptive functioning scale in the friend (OR=7.9 95% CI [1.67-37.43], p=0.02), and professional (OR=4.13 95% CI [1.33-12.83], p=0.007) areas. The ASR total problem score was also higher in patients (OR=5.12 95% CI [2.55-10.27], p<0.0001). This was confirmed by the adaptive profile evaluated by relatives (ACBL) (OR=7.57 95% CI [2.41-23.74], p=0.0005).

**Table 2.** Socio-demographic and Health related outcomes in patients with narcolepsy type 1 and controls. Comparisons between patients (questionnaires) and controls are adjusted for age and sex (last column, significant results in bold). BMI: Body Mass Index, HPB: Elevated Blood Pressure, ADHD: Attention Deficit with HyperActivity)

| Variable | Patients<br>(N=235) | Controls<br>(N=166) | adjusted p-value and OR [CI<br>95%] |
| --- | --- | --- | --- |
| **Marital Status, N (%)** |  |  | **p=0.004** |
| Single | 130 (55.8%) | 54 (32.5%) | 1 |
| Union | 88 (37.8%) | 102 (61.5%) | **0.38 [0.22-0.66]** |
| Divorced | 12 (5.1%) | 7 (4.2%) | 0.85 [0.29-2.56] |
| Widowed | 3 (1.3%) | 3 (1.8%) | 0.45 [0.08-2.47] |
| **Having children (yes), N(%)** | 95 (40.4%) | 102 (61.5%) | **p=0.006**<br>**0.56 [0.26-0.80]** |
| **BMI, N (%)** |  |  | **p<0.0001** |
| Underweight (<18.5 kg/m<sup>2</sup>)<br>or Normal (18.5-24.9 kg/m<sup>2</sup>) | 90 (38.9%) | 94 (57.0%) | 1 |
| Overweight (25-29.9 kg/m<sup>2</sup>) | 78 (33.8%) | 59 (35.7%) | 1.51 [0.94-2.41] |
| Obesity (>30kg/m<sup>2</sup>) | 63 (27.3%) | 12 (7.3%) | **6.12 [3.03-12.36]** |
| **Medical Past, N (%)** |  |  |  |
| HPB | 21(9.0%) | 12 (7.2%) | p=0.1 1.91 [0.84-4.36] |
| Diabetes | 8 (3.4%) | 5 (3.0%) | p=0.8 1.16 [0.35-3.83] |
| Hypercholesterolemia | 20 (8.6%) | 6 (3.6%) | **p=0.006 4.11 [0.50-11.422]** |
| Dysthyroid | 15 (6.5%) | 8 (4.8%) | p=0.3 1.65 [0.67-4.07] |
| Obesity surgery | 14 (6.0%) | 2 (1.2%) | **p=0.02 6.01 [1.32-27.35]** |
| ADHD | 8 (3.5%) | 0 (0.0%) | **p=0.02** |
| Depression | 58 (25.1%) | 30 (18.1%) | **p=0.02 3.06 [1.16-8.10]** |
| Treatment for depression | 24 (10.3%) | 12 (7.2%) | p=0.1 1.89 [0.89-3.99] |
| Treatment for anxiety | 12 (5.2%) | 7 (4.2%) | p=0.1 1.61 [0.60-4.30] |
| **Consumptions, N (%)** |  |  |  |
| Caffeinated drinks | 201 (86.3%) | 146 (88.0%) | p=0.5 0.82 [0.45-1.51] |
| Psychostimulants (apart<br>from medical prescription:<br>amphetamine, ecstasy etc.) | 17 (7.3%) | 6 (3.7%) | p=0.2 1.93 [0.73-5.10] |
| Tobacco |  |  | **p=0.03** |
| Current consumption | 75 (32.3%) | 33 (20.1%) | **1.98 [1.18-3.30]** |
| Past consumption | 47 (20.3%) | 40 (24.4%) | 1.25 [0.72-2.16] |
| Never | 110 (47.4%) | 91 (55.5%) | 1 |
| Cannabis consumption in<br>the last 6 months | 34 (14.6%) | 15 (9.1%) | p=0.2 1.5 [0.78-2.91] |
| Alcohol (AUDIT-C positive:<br>≥4 in men and ≥3 in women) | 69 (29.9%) | 50 (30.5%) | p=0.4 1.29 [0.75-2.22] |
| **Regular Physical activity, N**<br>**(%)** | 128 (55.2%) | 110 (67.1%) | **p=0.01 0.57 [0.38-0.88]** |
| **Enough leisure time,**<br>**« yes », N (%)** | 102 (47.7%) | 92 (57.5%) | **p=0.04 0.63 [0.41-0.97]** |

### Academic outcomes

The distribution of diploma obtained in patients and controls is presented in Figure 1. A significant difference (p=0.04) was found between patients from the registry and those who answered the questionnaires, with an overall lower level of graduation in the former group. No difference (p=0.5) was observed between patients and controls (questionnaires), nor between the three groups of patients according to age at disease onset and diagnosis. No difference was observed for the proportion of participants reporting at least one grade repetition throughout their schooling (48.9% in patients vs 42.7% in controls, p=0.2). The qualification obtained corresponded to the first choice of the vast majority of participants (82.9% vs 87.8%, p=0.1). However, patients reported more interruptions in academic progress (20% vs 8.5%, OR=2.63 95% CI [1.38-5.02], p=0.004), and qualitative differences in academic progress perception were observed; academic performance was considered as difficult in 59.0% of patients vs 23.8% of controls (OR=4.30 95% CI [2.72-6.79], p<0.0001), and patients reported more attention problems, absenteeism, and lateness in the morning (Table 3). This was more pronounced in patients with disease onset in childhood than in adulthood. In 76% of cases, school staff/colleagues were informed of the disease (mainly the school director, teachers, the school physician/nurse, and friends). Patients with childhood disease-onset benefited from specific support in 59.8% of cases when the narcolepsy was diagnosed vs 9.8% when it was not the case (p<0.0001). Specific support provided by schools consisted mostly of extra time for examinations (87.2%) and the possibility of taking naps at school (9.0%). They were mainly suggested (83.5%) by the sleep physician.

**Figure 1.**
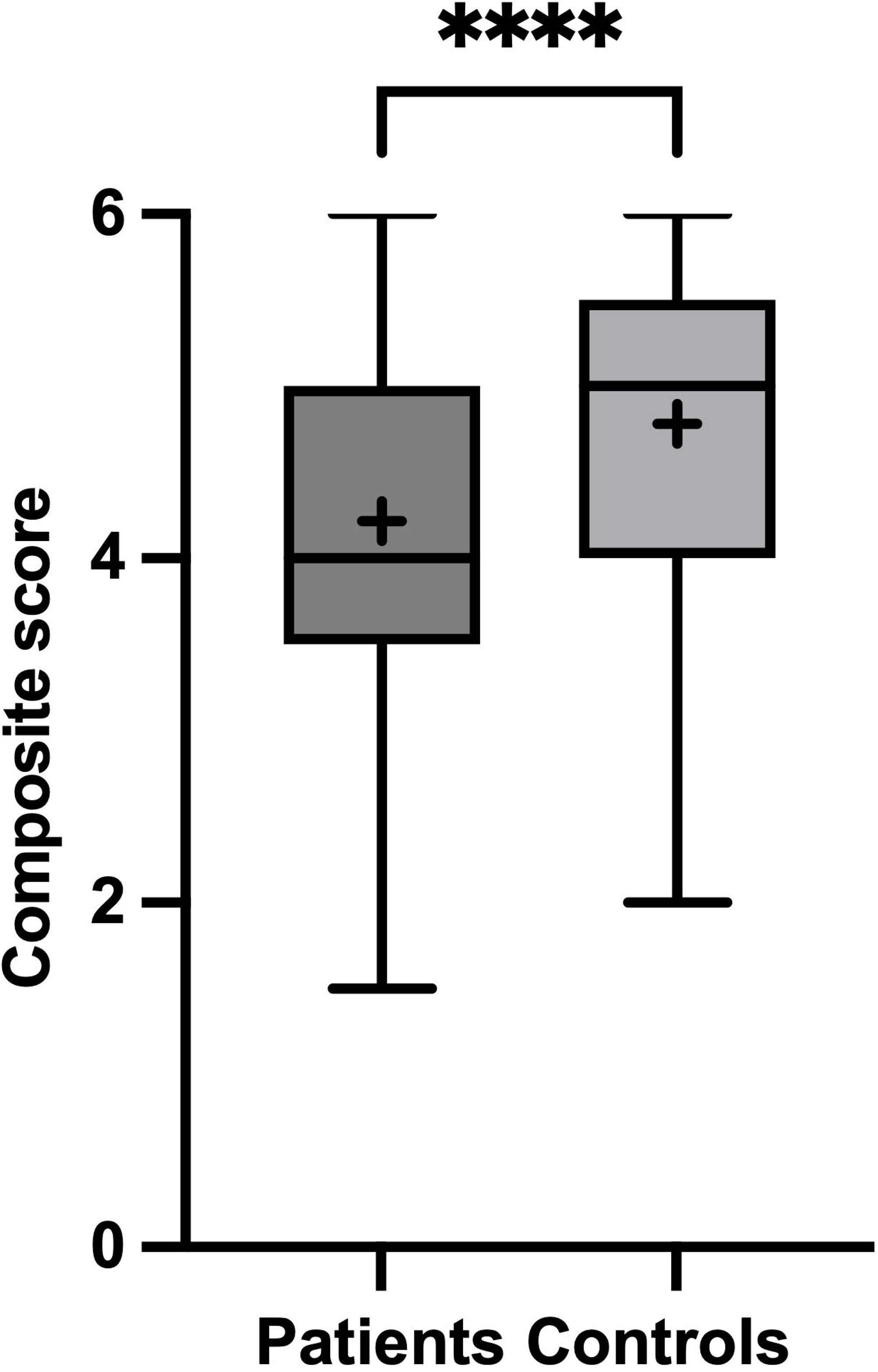
Distribution of the highest graduation obtained in patients with narcolepsy type 1 and controls. I : Graduate (Master 2) or doctorate; II: Bachelor - Master 1 (undergraduate) III : Advanced Technician Certificate IV: High School Graduation V: Certificate of Professional Competence VI: Middle School Level Missing data A. In all participants: Patients from registry (1164, missing data in 508), patients from questionnaire (235, missing data in 3) and controls (166, missing data in 3). A lower level of graduation is observed in patients from the registry (p=0.04) vs in those who answered the questionnaire. No significant difference (p=0.5) is observed between patients from questionnaire and controls. B. In NT1 patients (questionnaires) according to age at disease onset and age at diagnosis. No significant difference is observed after adjustement for age.

**Table 3.**
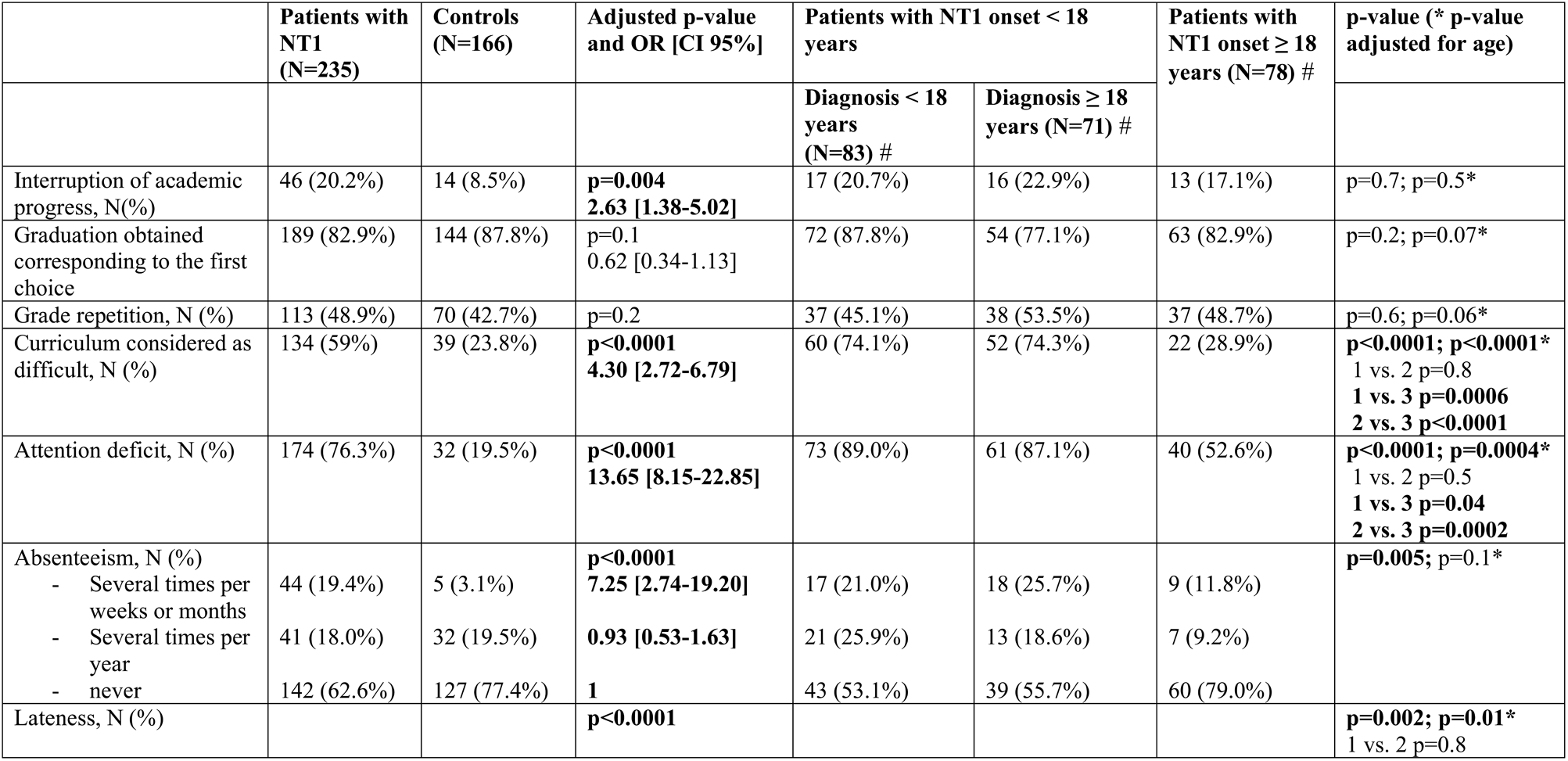
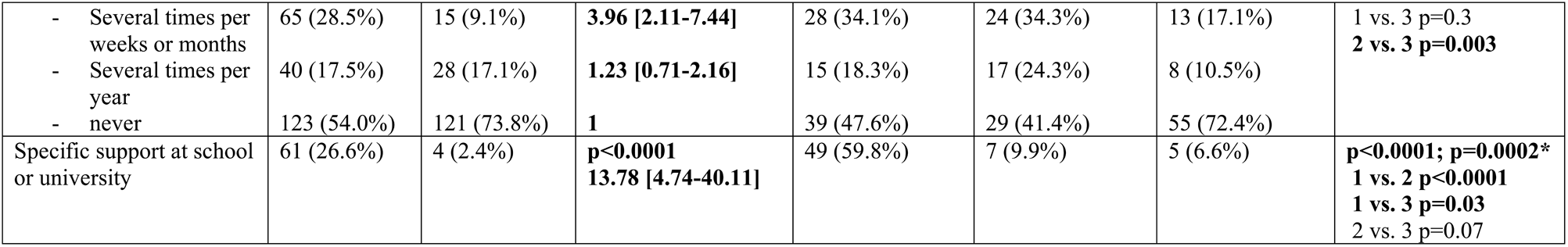
Academic path in patients with narcolepsy and controls. #note that the sum of the number of patients in the 3 groups is 232, as data bout age at onset and age at diagnosis were missing for 3 patients. Comparisons between the 2 (patients/controls) or 3 (patients according to age at disease and diagnosis onset) groups: significant results in bold

### Professional situation

No difference was observed in the distribution of parents’ SPC between patients (questionnaire) and controls. Regarding professional status, 69.5% of patients (questionnaires) were employed at the time of the study versus 77.0% of controls (p=0.2) (Figure 2). A significant difference (p=0.0001) was found between patients from the registry (53.4% of employement) and those from questionnaires, with overall more inactive people (20.7% vs 12.3%) and students/trainees (19.1% vs 11.4%) in the former group. SPC distributions did not differ between patients from questionnaires and controls (p=0.2). No difference was found either for the presence of unemployment periods during their career nor for the number of changes in position (Table 4). However, the reasons for changes differed between the two groups, with less “desired” changes (e.g. promotion) and more “unwanted” (e.g. dismissal) in patients (p=0.005). Part-time work was more prevalent among patients than controls (38.9% vs 25.3%, OR=1.91 95% CI [1.20-3.04], p=0.007). No difference was observed in work schedules but more patients reported working from home. Regarding adaptative behaviors, more patients than controls reported napping occasionally or frequently (respectively 24.1% vs 6.3% and 27.3% vs 0.6%, p<0.0001). Specific support was provided at work for 21% of patients, consisting of schedule adaptation (52.2%), providing a time and a place for naps (58.7%), or allowing flexible organization (56.5%) and work from home (32.6%). Patients reported being late at work more often than controls with 45.6% (vs 22.8%, p<0.0001) being late several times per year, months or week. The employer was informed about the disease in 50.7% of the patients (61.5% for occupational physicians and 76.5% for colleagues). The majority (51.6%) of the patients reported feeling accepted as a narcoleptic person at the workplace, but 16% felt misunderstood and thought that they were considered lazy. Sleepiness (59.2%) and attention deficit (23.9%) were considered the most disabling symptoms, followed by cataplexy (8.5%) and sleep inertia (7.5%). Many patients had developed coping strategies to adapt to their disease, such as hiding away to nap (50.7%), hyperactive behavior (40.7%) or avoiding meetings (41.3%), promotions (18.3%), and examinations (22.5%). Effort-reward imbalance (Siegrist score) was increased in patients vs controls (ratio effort/reward 0.65±0.57 vs 0.54±0.26, OR=2.28 95% CI [1.20-4.33], p=0.01), essentially due to significant lower reward (p=0.0002). Patients reported being disturbed by overtime work and being pressed for time, felt their job stability was under threat, with fewer prospects for promotion, and scored higher for sleep disturbances due to worries about procrastination.

**Figure 2.**
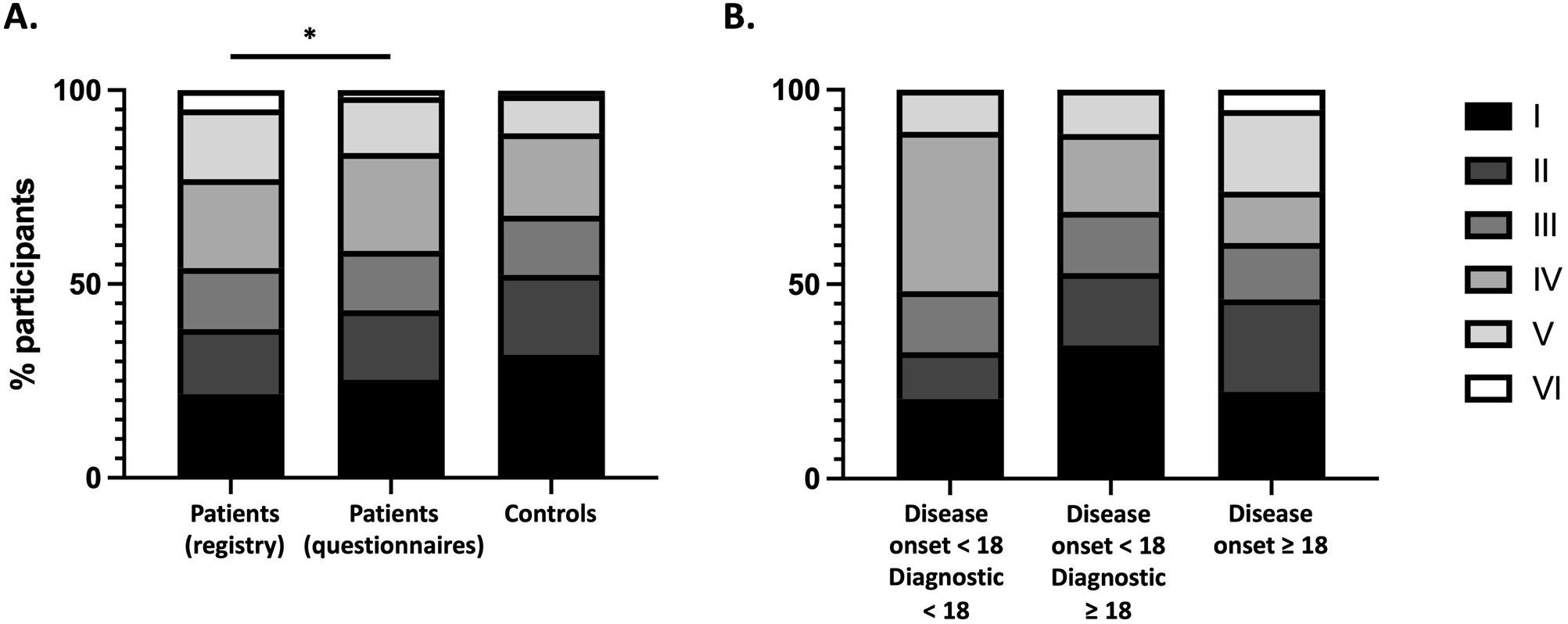
Distribution of professionnal status categories in patients with narcolepsy type 1 and controls. Status categories: Employed (Fixed-term contract, indefinite term contract, civil servant, permanent employee, temporary worker, self-employed…); Unemployed (Job loss / Housewife/Househusband / Disability); Retired; Student/Trainee A. In all participants: Patients from registry (N=1164, missing data in 251), Patients from questionnaires (N=235, missing data in 16), Controls (166, missing data in 5). A significant difference is observed between patient from the registry and thoses from questionnnaires (p=0.0001) whereas no difference is found between patients from questionnaires and controls, B. In NT1 patients (questionnaires) according to age at disease onset and age at diagnosis. No significant difference is observed after adjustement for age.

**Table 4.**
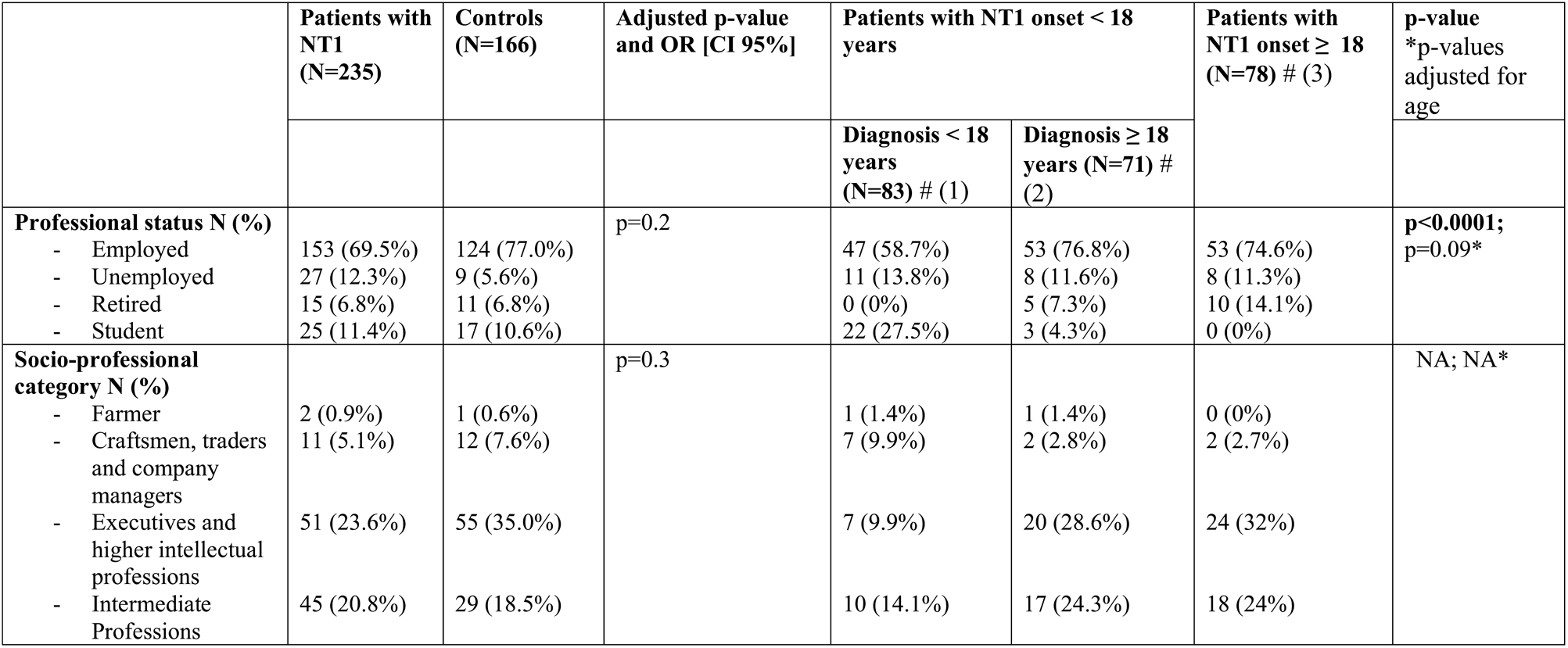
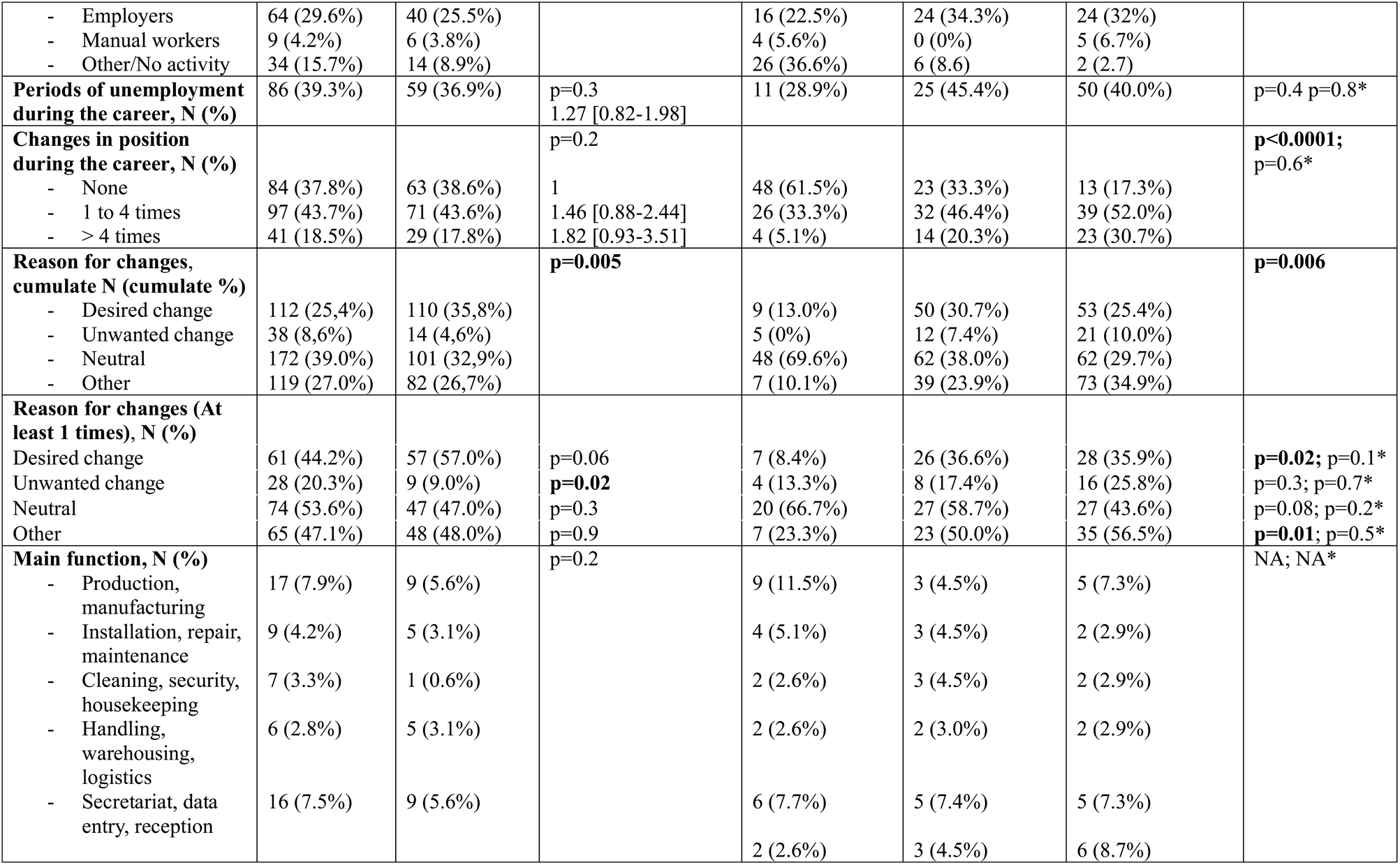
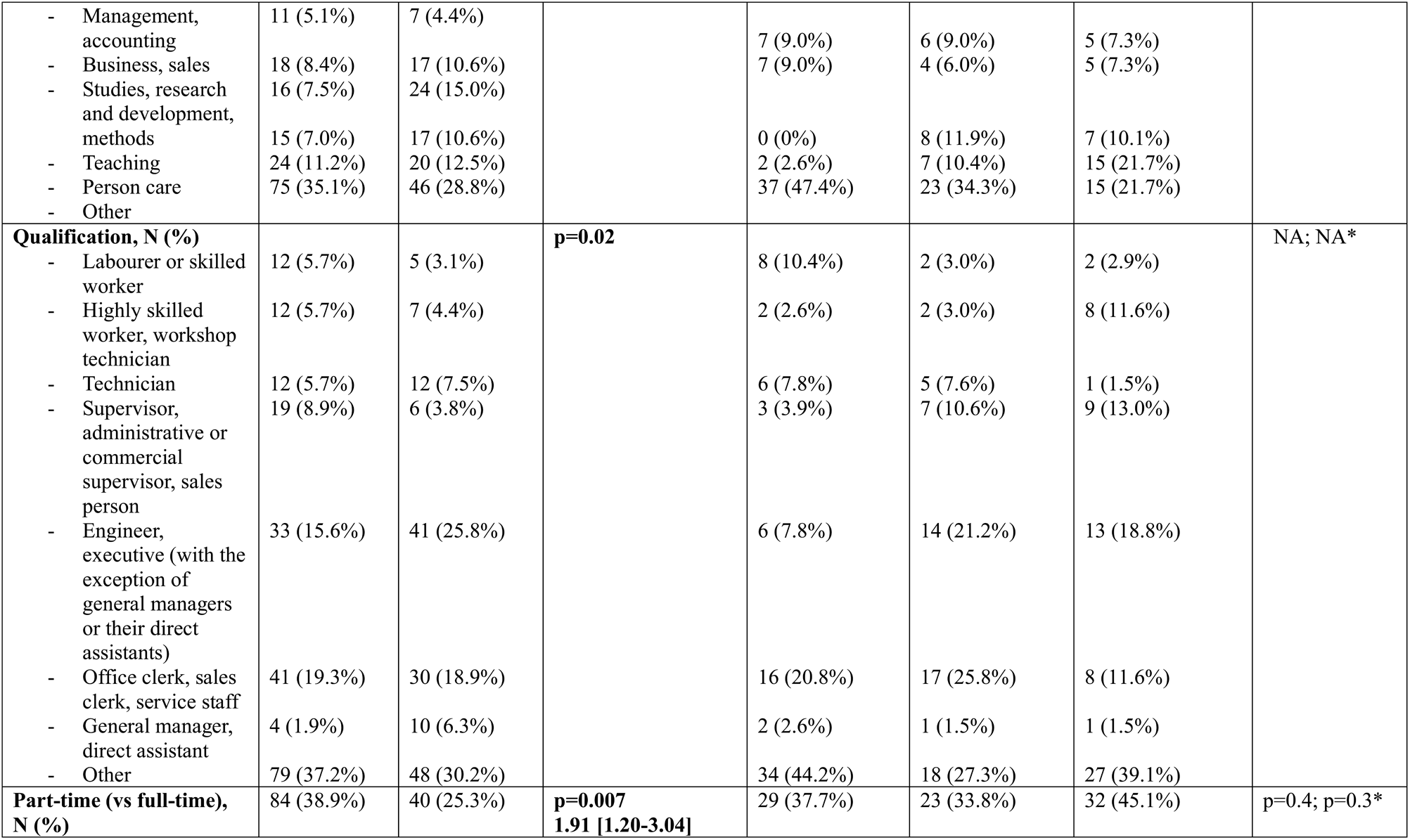
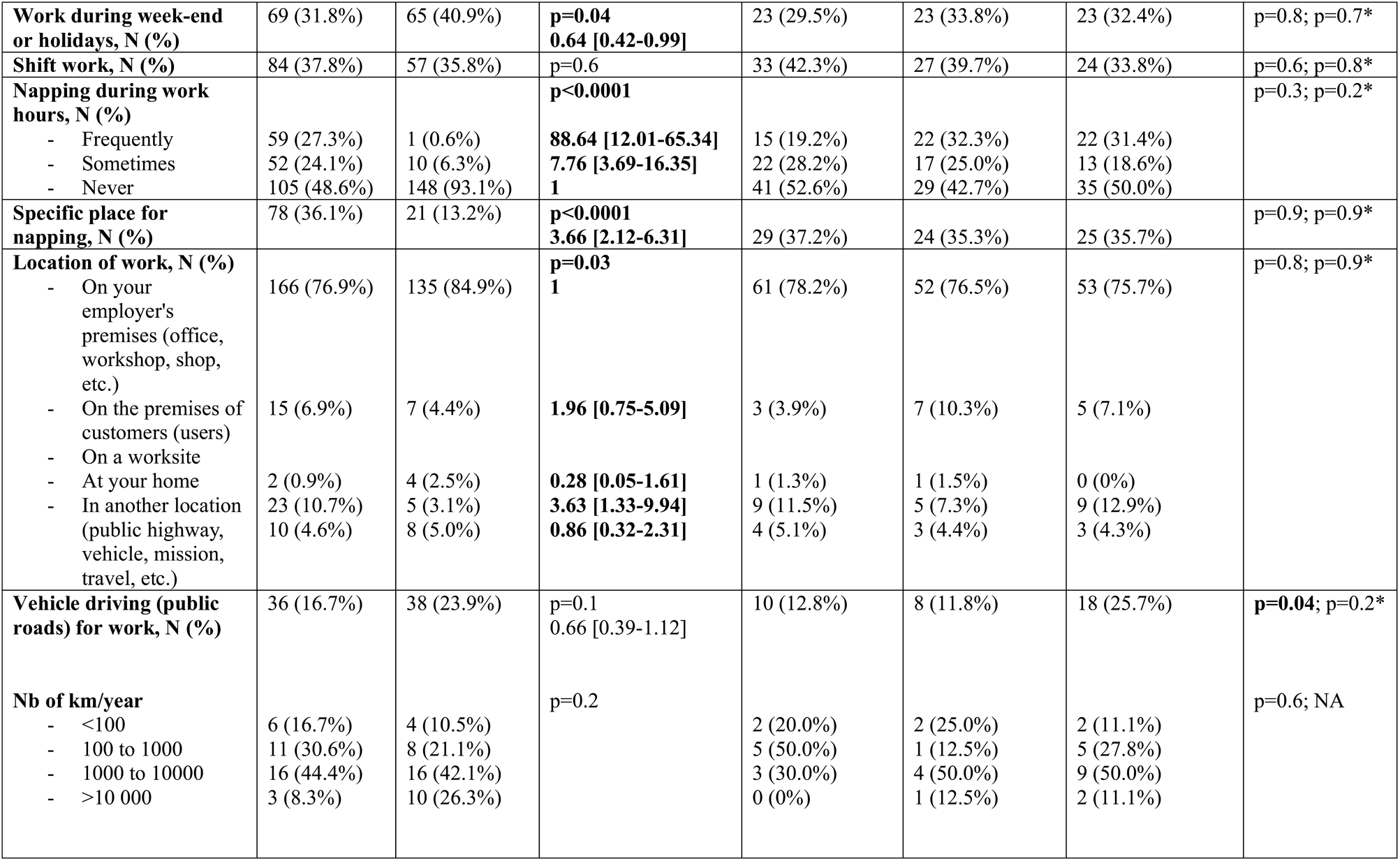
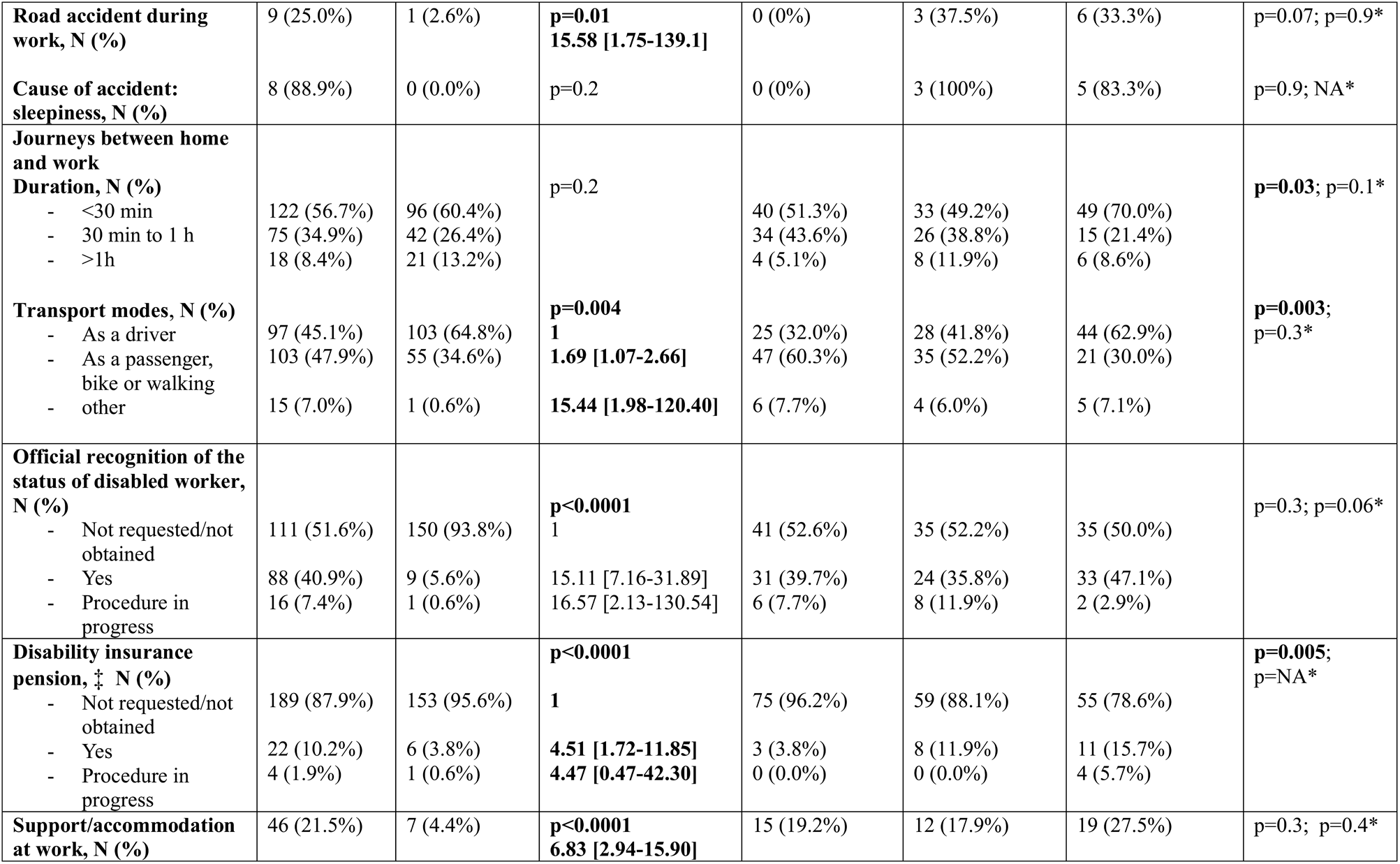
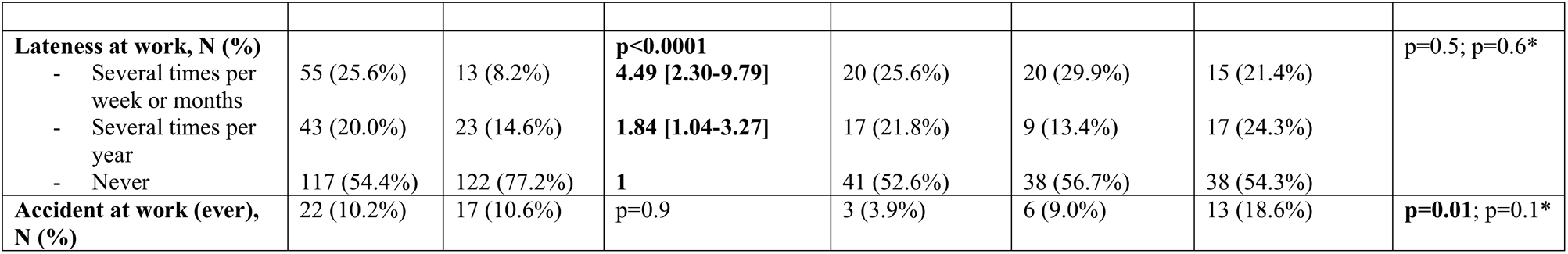
Professional path in patients with narcolepsy and controls. #note that the sum of the number of patients in the 3 groups is 232, as data bout age at onset and age at diagnosis were missing for 3 patients. Comparisons between the 2 (patients/controls) or 3 (patients according to age at disease and diagnosis onset) groups: significant results in bold ‡ Disability insurance pension (“invalidity”), N=22: 13 category 1 (part-time work possible) and 9 category 2 or 3 (no work) Analyses are adjusted for age and sex (patients vs controls) and for age (patients subcategories) NA: not applicable (low sample) Reasons for end of contract: desired change (=promotion, voluntary quit), undesired change (=dismissal), neutral (=end of contract), others

Almost half of the patients benefited from an official recognition of disability and 10.2% received invalidity benefits (with disability either preventing work [4.2%] or allowing part-time activity [6.0%]). The official recognition of disability was reported by most severe cases with higher NSS score (p=0.01), higher BDI-II score (p=0.0005), lower EQ5D-VAS score (p=0.007), and more attentional disorders on ASR (p=0.009). Patients who received invalidity benefits were older (p<0.0001), had lower EQ5D-VAS score (p=0.007), more adaptative behavioral dysfunction (p=0.04) and depression/anxiety symptoms (p=0.007) as well attention disorders (p=0.003) on the ASR scale.

The distribution of professional status differed between the 3 groups of patients, but this difference did not remain significant after adjustment for age, as for all work-related outcomes detailed in Table 4.

### Prognostic factors

In order to explore factors associated with a more favourable professional progress, a composite score was used as detailed in the method section.^19^ This score was calculated in patients and controls (questionnaires), excluding students and retired people. It was significantly lower in patients than in controls (4.2±1.1 (N=179) vs 4.8±0.9 (N=133), p<0.0001) (Figure 3). Univariate analysis highlighted that several factors were associated with the prognosis: obesity (r=-0.17, p=0.007), sleepiness (r=-0.06, p=0.004), depression (r=-0.37, p<0.0001), quality of life (r=0.01, p=0.004), clinical/limit range total score at the adaptative functioning scale (r=0.31, p=0.002), attention disorders (r=0.41, p=0.0002), effort-reward imbalance (r=-0.43, p=0.002), being narcoleptic during academic path (r=0.52, p=0.003). On the contrary, age (p=0.07), sex (p=0.9), NSS (p=0.1), age at diagnosis (p=0.2) and disease onset (p=0.1), diagnosis delay (p=0.4), patient group (p=0.1), treatments (p>0.05 for all drugs and number of drugs), official recognition of the status of disabled student/worker (p=0.4 and p=0.08) and specific support at school/work (p=0.6 and p=0.08) were not associated with prognosis. The multivariate matched analysis identified three remaining significant factors; depression and attention disorders being associated with a worse prognosis (r=-0.40, p<0.0001 and r=0.26, p=0.03) and being narcoleptic during schooling, which was associated with a better prognosis (r=0.44, p=0.02).

**Figure 3:**
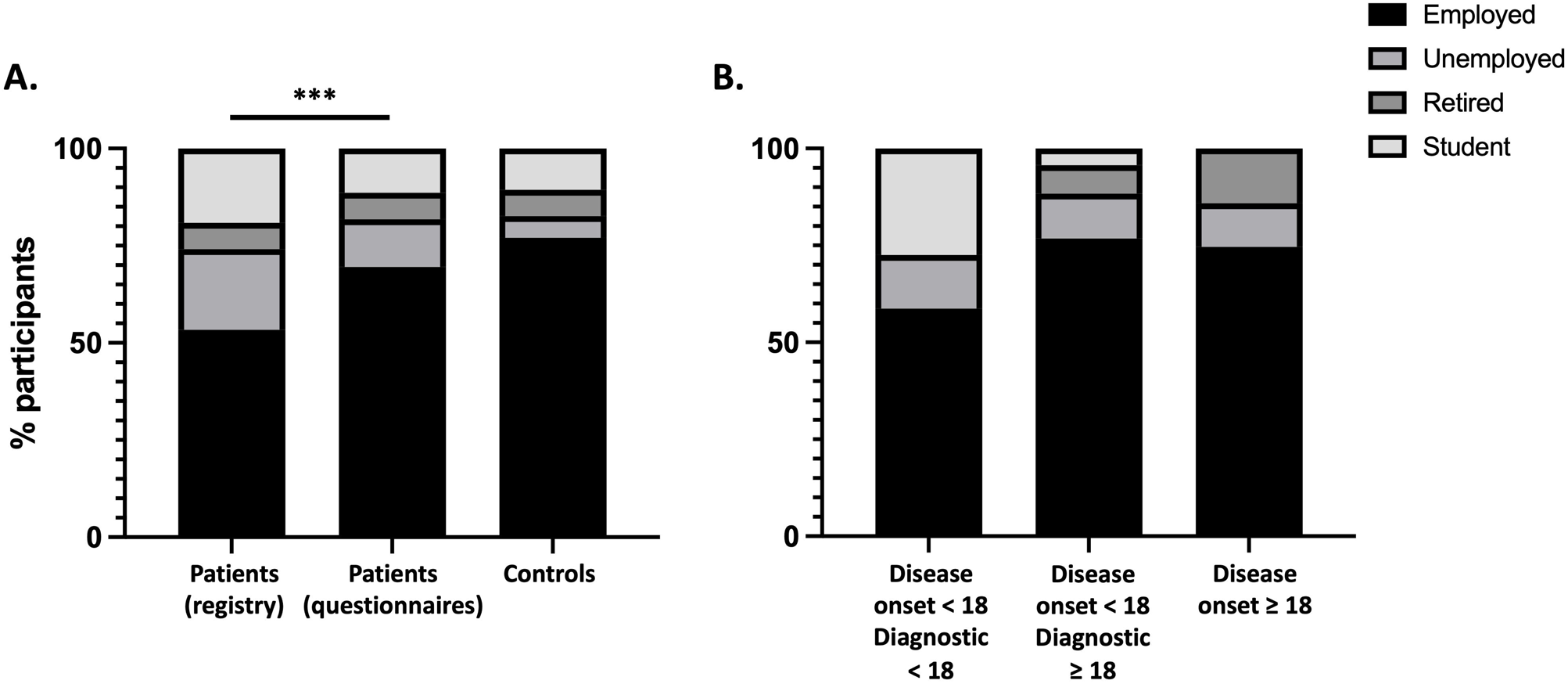
Composite prognosis score in patients with NT1 and controls. The prognosis score includes the level of graduation, the cumulate duration of periods of unemployment, the number of job changes due to the disease or dismissal, the current professional status, the frequency of being late for work and the number of sick leave days during the past years; it ranges from 0 (lowest prognosis) to 6 (best prognosis). A better prognosis is observed in controls than in patients with NT1 (questionnaires) ((4.2±1.1 vs 4.8±0.9, p<0.0001)).

### Economic costs

The mean monthly income based on the SPC was 2603±1097 € in patients and 3125±1164 € in controls (p<0.001) with the assumption of full-time work, and 2316±1103 € vs 3023±1155 € (p<0.001) considering part-time work. No significant difference was observed between the two groups for the proportion of participants with at least one sick leave (26.1% vs 21%, p=0.094) and the average duration of work stoppage (15.5±51.4 vs 8.7±37.3 days, p=0.096). The annual loss of income due to missed days was 736±2922 € for patients and 473±1788 € for controls (p<0.001). From the societal perspective, the average annual cost of productivity loss was 1680±5755 € for patients and 1004±4346 € for controls based on GDP (p=0.098) and 2240±8587€ vs 1398±5814 € based on SPC (p=0.134). Results of sensitivity analyses are summarized in Supplementary Table 3.

## Discussion

The Narcoscol-Narcovitae study aimed to investigate the educational and occupational trajectories of patients with NT1 in France. We observed (i) no difference in diploma and grade repetitions in patients vs controls but academic progress was considered more difficult by patients (ii) no difference in employment rate and SPC but the effort-reward imbalance was increased in patients (iii) a significant association between impaired professional prognosis and depression and attention disorders, while being narcoleptic during schooling allowed a better prognosis.

In line with the Narcowork pilot study, no difference was found for the level of graduation in patients vs controls.^19^ Notably, the proportion of subjects with post-high school diplomas in patients from the registry (54.1%), patients from questionnaires (58.5%) and controls (67.6%) was higher than values reported in the 35-44 years general population in France in 2020 (50.9% in women and 42.4% in men) (National Institute of Statistics and Economic Studies). However, interrupted academic progress, absenteeism and lateness were more frequent in patients with childhood onset disease. This was the case even when the diagnosis had been established in childhood and despite specific support at school, keeping in mind that such support may only be offered in the most difficult situations.^37^ Comorbidities are likely to contribute to the disability, such as attentional disorders reported by almost 90% of patients in our study, or other psycho-behavioral symptoms.^8, 38^ Importantly, it is possible that the achievement of academic “success” in terms of graduation may be at the expense of other areas associated with well-being and quality of life such as after-school activities or friendships.^38, 39^

Our findings regarding career outcomes echo those observed for the academic curriculum. Although the overall impact on employment rate was not significant, several indicators reflect the difficulties experienced by patients: part-time work, unwanted changes in position, fewer prospects of promotion, and fear of job loss. Their income was lower, and the cost due to sick leave days was high for patients as previously reported.^4, 20–22, 26^ This lack of professional recognition and valuing resulted in an effort-reward imbalance, as patients living with NT1 reported investing most of their energy in work, possibly to the detriment of their personal lives. Thus, as highlighted by previous studies, patients were less likely to be in a couple, had fewer children, and expressed more difficulty in managing daily activities and engaging in leisure activities.^18, 26, 29, 40^ However, patients described relatively high adaptations such as official recognition of the status, napping at work and, flexible working organization. Such adaptations are likely to relieve the pressure exerted on patients, as suggested by reports of positive consequences of remote work/school during the COVID 19 lockdown.^41, 42^

Psycho-behavioral disorders are frequent and affect multiple dimensions in NT1, although their determinism (reaction to symptoms such as somnolence or cataplexy or to the disability and stigmatization associated with the disease, or direct involvement of orexinergic dysfunction) is not fully understood.^5, 6, 12, 13, 15, 43, 44^ We observed that depression was strongly associated with professionnal prognosis with a possible bidirectional link: depression preventing patients from performing well at work and psychopathological risk being aggravated by academic/professional failure. Attention disorders were also a significant predictor, which suggests that in treated patients, cognitive disorders have a greater impact on daily functioning than sleep-related symptoms, although sleepiness might impact other aspects such as work productivity.^27^ Being narcoleptic during schooling was associated with a better professionnal prognosis. For patients diagnosed in childhood, schooling may offer a period of adjustment, potentially useful for the rest of their academic and professional lives (e.g. in terms of coping strategies or in career choices), and may contribute to resilience, which is negatively correlated with depression in patients with NT1.^18, 45, 46^ The diagnosis delay was not a determining factor, possibly as many factors, differentially influencing the prognosis, are associated with a short delay; younger age at onset, higher symptom severity (especially cataplexy), year of the diagnosis, and social environment associated with inequities in specialized medical care access.^47, 48^ Medication was not a significant prognosis factor as already reported for quality of life, possibly as most patients were treated.^29^ However, some studies suggest that appropriate treatment may favorably impact educational path and work productivity.^12, 16, 49^ Last, the official recognition of the disability, aiming to facilitate support, was not associated with a better prognosis. In the context of a cross-sectional study, it is impossible to decipher the real impact of this measure, which was probably requested by the most disabled patients.

The main limitation of our study is the low response rate to questionnaires, with likely a selection of the least severe patients. In addition, the gender imbalance, which does not reflect the actual sex ratio of the disease, is that frequently observed in health research surveys where women are more willing to participate.^50^ We anticipated this limitation by filling a registry with a minimal dataset, which enabled us to collect the main outcomes for the majority of patients seen in the centers during the inclusion period; this confirmed that the employment rate and graduation level were lower in non-respondent than in respondent patients. This methodological bias may explain why studies based on registries have reported poorer prognosis than works based on questionnaires.^20, 26^ Importantly, one must also consider the hidden part of the iceberg, i.e. patients who are not diagnosed or who do not seek care either because they are less severely affected or for socio-economic reasons.^48^ Overall, our study may have underestimated patients’ disability; however our methodology provided extremely accurate and extensive data, enabling us to provide an in-depth picture of patients’ educational and professional situations. In addition, the composition of the control group, consisting of two acquaintances of the patients, helped to minimize bias arising from variations in social background. Finally, our results about medico-economic outcomes may not be generalized as financial support for people with disability may vary from one country to another.

As a conclusion, our findings suggest that most patients with NT1 treated in specialized centers manage to achieve their career goals, but that the price paid in terms of daily efforts, concessions on other aspects of their lives and psychological burden is high in relation to the reward obtained. More than sleep-related symptoms, co-morbidities play a critical role in the educational and professional prognosis of patients. This reinforces the need for a comprehensive care of patients suffering from narcolepsy, including psycho-cognitivo-social dimensions. Future prospective studies are needed to assess which measures (pharmacological or non-pharmacological treatment, psychological support, development of psychosocial skills, adaptation of the environment…) are likely to reduce the academic and professional burden associated with the disease.

## References

1. Mahoney CE, Cogswell A, Koralnik IJ, Scammell TE. The neurobiological basis of narcolepsy. Nat Rev Neurosci 2019;20:83–93.

2. Barateau L, Pizza F, Chenini S, Peter-Derex L, Dauvilliers Y. Narcolepsies, update in 2023. Rev Neurol (Paris) 2023.

3. Raggi A, Plazzi G, Ferri R. Health-Related Quality of Life in Patients With Narcolepsy: A Review of the Literature. J Nerv Ment Dis 2019;207:84–99.

4. Flores NM, Villa KF, Black J, Chervin RD, Witt EA. The Humanistic and Economic Burden of Narcolepsy. J Clin Sleep Med 2016;12:401–407.

5. Filardi M, D’Anselmo A, Agnoli S, et al. Cognitive dysfunction in central disorders of hypersomnolence: A systematic review. Sleep Med Rev 2021;59:101510.

6. Ruoff CM, Reaven NL, Funk SE, et al. High Rates of Psychiatric Comorbidity in Narcolepsy: Findings From the Burden of Narcolepsy Disease (BOND) Study of 9,312 Patients in the United States. J Clin Psychiatry 2017;78:171–176.

7. Gudka S, Haynes E, Scotney J, et al. Narcolepsy: Comorbidities, complexities and future directions. Sleep Med Rev 2022;65:101669.

8. Blackwell JE, Alammar HA, Weighall AR, Kellar I, Nash HM. A systematic review of cognitive function and psychosocial well-being in school-age children with narcolepsy. Sleep Medicine Reviews 2017;34:82–93.

9. Plazzi G, Clawges HM, Owens JA. Clinical Characteristics and Burden of Illness in Pediatric Patients with Narcolepsy. Pediatr Neurol 2018;85:21–32.

10. Thieux M, Zhang M, Marcastel A, et al. Intellectual Abilities of Children with Narcolepsy. J Clin Med 2020;9.

11. Inocente CO, Gustin M-P, Lavault S, et al. Quality of life in children with narcolepsy. CNS Neurosci Ther 2014;20:763–771.

12. Rocca FL, Finotti E, Pizza F, et al. Psychosocial Profile and Quality of Life in Children With Type 1 Narcolepsy: A Case-Control Study. Sleep 2016;39:1389–1398.

13. Stores G, Montgomery P, Wiggs L. The psychosocial problems of children with narcolepsy and those with excessive daytime sleepiness of uncertain origin. Pediatrics 2006;118:e1116–1123.

14. Broughton R, Ghanem Q, Hishikawa Y, Sugita Y, Nevsimalova S, Roth B. Life effects of narcolepsy in 180 patients from North America, Asia and Europe compared to matched controls. Can J Neurol Sci 1981;8:299–304.

15. Lecendreux M, Lavault S, Lopez R, et al. Attention-Deficit/Hyperactivity Disorder (ADHD) Symptoms in Pediatric Narcolepsy: A Cross-Sectional Study. Sleep 2015;38:1285–1295.

16. Aran A, Einen M, Lin L, Plazzi G, Nishino S, Mignot E. Clinical and therapeutic aspects of childhood narcolepsy-cataplexy: a retrospective study of 51 children. Sleep 2010;33:1457–1464.

17. Dias Costa F, Barreto MI, Clemente V, Vasconcelos M, Estêvão MH, Madureira N. Narcolepsy in pediatric age - Experience of a tertiary pediatric hospital. Sleep Sci 2014;7:53–58.

18. Ingravallo F, Gnucci V, Pizza F, et al. The burden of narcolepsy with cataplexy: how disease history and clinical features influence socio-economic outcomes. Sleep medicine 2012;13:1293–1300.

19. White M, Charbotel B, Fort E, et al. Academic and professional paths of narcoleptic patients: the Narcowork study. Sleep Med 2020;65:96–104.

20. Jennum P, Ibsen R, Kjellberg J. Long-term health and socioeconomic consequences of childhood and adolescent-onset of narcolepsy. Sleep Medicine 2020;67:23–27.

21. Dodel R, Peter H, Walbert T, et al. The socioeconomic impact of narcolepsy. Sleep 2004;27:1123–1128.

22. Jennum P, Knudsen S, Kjellberg J. The economic consequences of narcolepsy. J Clin Sleep Med 2009;5:240–245.

23. Ozaki A, Inoue Y, Hayashida K, et al. Quality of life in patients with narcolepsy with cataplexy, narcolepsy without cataplexy, and idiopathic hypersomnia without long sleep time: comparison between patients on psychostimulants, drug-naive patients and the general Japanese population. Sleep medicine 2012;13:200–206.

24. Broughton RJ, Guberman A, Roberts J. Comparison of the psychosocial effects of epilepsy and narcolepsy/cataplexy: a controlled study. Epilepsia 1984;25:423–433.

25. Taddei RN, Werth E, Poryazova R, Baumann CR, Valko PO. Diagnostic delay in narcolepsy type 1: combining the patients’ and the doctors’ perspectives. J Sleep Res 2016;25:709–715.

26. Jennum P, Ibsen R, Petersen ER, Knudsen S, Kjellberg J. Health, social, and economic consequences of narcolepsy: a controlled national study evaluating the societal effect on patients and their partners. Sleep Med 2012;13:1086–1093.

27. Bassi C, Biscarini F, Zenesini C, et al. Work productivity and activity impairment in patients with narcolepsy type 1. J Sleep Res 2023:e14087.

28. Dodel R, Peter H, Spottke A, et al. Health-related quality of life in patients with narcolepsy. Sleep medicine 2007;8:733–741.

29. Daniels E, King MA, Smith IE, Shneerson JM. Health-related quality of life in narcolepsy. J Sleep Res 2001;10:75–81.

30. American Academy of Sleep M. International Classification of Sleep Disorders, 3rd ed. (American A). IL: Darien.2014.

31. Johns MW. A new method for measuring daytime sleepiness: the Epworth sleepiness scale. Sleep 1991;14:540–545.

32. Beck AT, Steer RA, Brown GK. Manual for the Beck Depression Inventory-II. San Antonio, TX: Psychological Corporation.1996.

33. Dauvilliers Y, Beziat S, Pesenti C, et al. Measurement of narcolepsy symptoms: The Narcolepsy Severity Scale. Neurology 2017;88:1358–1365.

34. Siegrist J, Starke D, Chandola T, et al. The measurement of effort-reward imbalance at work: European comparisons. Soc Sci Med 2004;58:1483–1499.

35. EuroQol--a new facility for the measurement of health-related quality of life. Health Policy 1990;16:199–208.

36. Achenbach TM, Rescorla L. Manual for the ASEBA adult forms & profiles. Burlington, VT: University of Vermont, Research Center for Children, Youth …, 2003.

37. Janssens K, Amesz P, Nuvelstijn Y, et al. School Problems and School Support for Children with Narcolepsy: Parent, Teacher, and Child Reports. Int J Environ Res Public Health 2023;20.

38. Avis KT, Shen J, Weaver P, Schwebel DC. Psychosocial Characteristics of Children with Central Disorders of Hypersomnolence Versus Matched Healthy Children. J Clin Sleep Med 2015;11:1281–1288.

39. Zhou ES, Revette A, Heckler GK, Worhach J, Maski K, Owens JA. Building a deeper understanding of social relationship health in adolescents with narcolepsy disorder. J Clin Sleep Med 2023;19:491–498.

40. Davidson RD, Biddle K, Nassan M, Scammell TE, Zhou ES. The impact of narcolepsy on social relationships in young adults. J Clin Sleep Med 2022;18:2751–2761.

41. Nigam M, Hippolyte A, Dodet P, et al. Sleeping through a pandemic: impact of COVID-19-related restrictions on narcolepsy and idiopathic hypersomnia. J Clin Sleep Med 2022;18:255–263.

42. Zhao M, Zhang B, Tang J, Zhang X. The Impact of Sleep Pattern in School/Work Performance During the COVID-19 Home Quarantine in Patients With Narcolepsy. Front Neurol 2022;13:849804.

43. Kapella MC, Berger BE, Vern BA, Vispute S, Prasad B, Carley DW. Health-related stigma as a determinant of functioning in young adults with narcolepsy. PLoS One 2015;10:e0122478.

44. Maski K, Steinhart E, Williams D, et al. Listening to the Patient Voice in Narcolepsy: Diagnostic Delay, Disease Burden, and Treatment Efficacy. J Clin Sleep Med 2017;13:419–425.

45. D’Alterio A, Menchetti M, Zenesini C, et al. Resilience and its correlates in patients with narcolepsy type 1. J Clin Sleep Med 2023;19:719–726.

46. Vignatelli L, D’Alessandro R, Mosconi P, et al. Health-related quality of life in Italian patients with narcolepsy: the SF-36 health survey. Sleep medicine 2004;5:467–475.

47. Luca G, Haba-Rubio J, Dauvilliers Y, et al. Clinical, polysomnographic and genome-wide association analyses of narcolepsy with cataplexy: a European Narcolepsy Network study. J Sleep Res 2013;22:482–495.

48. Kim TJ, Vonneilich N, Lüdecke D, von dem Knesebeck O. Income, financial barriers to health care and public health expenditure: A multilevel analysis of 28 countries. Soc Sci Med 2017;176:158–165.

49. Emsellem HA, Thorpy MJ, Lammers GJ, et al. Measures of functional outcomes, work productivity, and quality of life from a randomized, phase 3 study of solriamfetol in participants with narcolepsy. Sleep Med 2020;67:128–136.

50. Glass DC, Kelsall HL, Slegers C, et al. A telephone survey of factors affecting willingness to participate in health research surveys. BMC Public Health 2015;15:1017.

