## Supplementary tables for "Academic and socio-professional trajectories in narcolepsy type 1: the NARCOSCOL-NARCOVITAE multicentric study"

### Supplementary table 1. Questionnaire about academic and occupational path

This customized questionnaire was built by a sleep physician, an occupational physician, a neuropsychologist, a psychiatrist and patients' association "ANC" (Association Narcolepsy-Cataplexy and hypersomnia)

| **Domain** | **Questions** |
|---|---|
| Demographic information | Age; sex |
| Medial past and treatments | High blood pressure; diabetes; hypercholesterolemia; dysthyroid; Attention Deficit with Hyperactivity; obesity surgery; depression; other conditions<br>Treatments<br>Weight; height |
| Use of substances<br>- Psychostimulants<br>- Cannabis<br>- Alcohol<br>- Tobacco | Caffeinated drinks and psychostimulant substances without prescription |
| Narcolepsy characteristics | Age at onset; age at diagnosis; symptoms (excessive daytime sleepiness, cataplexy, disrupted nighttime sleep, sleep-related hallucinations, sleep paralysis, sleep inertia); past and present treatments |
| Academic path | Last diploma obtained; interruption/reorientation during schooling; attention issues; absenteeism; lateness in the morning; grade repetition; official recognition of disability; specific support/accommodation due to narcolepsy; information and attitude of family, friends and teaching staff |
| Physical activity and leisure activities | Regular practice of physical activity; Having enough time for leisure activity / friends |
| Family | Relationship/marital status; number of children Having enough time spent with family |
| Professional career | Socio-professional categories of parents; Current status (employed [Fixed-term contract, indefinite term contract, civil servant, permanent employee, temporary worker, self-employed…], unemployed [loss of job, housewife/househusband, disability limiting work], Retired, or Student/trainee); socio-professional category; work history (periods of unemployment, changes in positions and reasons for change; present position and work characteristics (work schedule, full/part-time position, transport, remote work, breaks etc.); official recognition of disability; specific support/accommodation at the workplace including naps; absenteeism; lateness; accidents at work; information and attitude of occupational physician/colleagues/employer |

**Supplementary table 2. Demographic characteristics and Questionnaires results in patients with narcolepsy type 1 according to age at onset and age at diagnosis**

#note that the sum of the number of patients in the 3 groups is 232, as data bout age at onset and age at diagnosis were missing for 3 patients.
Comparisons between the 3 groups: significant results in bold

| | **Narcolepsy onset < 18 years** | | **Narcolepsy onset ≥ 18 years (N=78) (group 3)** | p-value<br>p-value* adjusted for age (with post-hoc) |
|---|---|---|---|---|
| | **Diagnosis of narcolepsy < 18 years (N=83) (group 1)** | **Diagnosis of narcolepsy ≥ 18 years (N=71) (group 2)** | | |
| **Sex, Women** N (%) | 53 (63.9%) | 46 (64.8%) | 50 (64.1%) | p=0.9; p=0.7* |
| **Age**<br>Mean ± std<br>Median (Q1-Q3) | <br>25.5 ± 7.7<br>23 (20-27) | <br>37.9 ± 13.9<br>35 (26-48) | <br>46.3 ± 12.9<br>44.5 (36-42) | **p<0.0001** |
| **BMI,** N (%)<br>Underweight or Normal (< 25 kg/$m^2$)<br>Overweight (25-29.9 kg/$m^2$)<br>Obesity (>30kg/$m^2$) | <br>31 (38.3%)<br>25 (30.9%)<br>25 (30.9%) | <br>34 (47.9%)<br>19 (26.8%)<br>18 (25.3%) | <br>25 (32.9%)<br>33 (43.4%)<br>18 (23.7%) | p=0.2; p=0.2* |
| **Consumptions,** N (%)<br>Caffeinated drinks<br>Psychostimulants (apart from medical prescription: amphetamine, ecstasy etc.)<br>Tobacco<br>Current consumption<br>Past consumption<br>Never<br>Cannabis consumption in the last 6 months<br>Alcohol (AUDIT-C positive: ≥4 in men and ≥3 in women) | <br>73 (87.9%)<br>4 (4.8%)<br><br>19 (22.9%)<br>6 (7.2%)<br>58 (69.9%)<br>11 (13.3%)<br>25 (30.1%) | <br>58 (81.7%)<br>7 (9.9%)<br><br>22 (31.0%)<br>16 (22.5%)<br>33 (46.5%)<br>10 (14.1%)<br>22 (31.4%) | <br>69 (88.5%)<br>6 (7.7%)<br><br>33 (42.9%)<br>25 (32.5%)<br>19 (24.7%)<br>13 (16.7%)<br>22 (28.2%) | <br>p=0.4; p=0.3*<br>p=0.5; p=0.3*<br>**p=<0.0001**; p=0.06*<br><br><br><br>p=0.8; p=0.7*<br>p=0.9; p=0.9* |
| **Regular Physical activity,** N (%) | 44 (53.0%) | 49 (69.0%) | 35 (44.9%) | **p=0.01;p=0.01***<br>**1 vs 2 p=0.03**<br>1 vs 3 p=0.4<br>**2 vs 3 p=0.06** |
| **ESS**<br>Mean ± sd<br>Median (Q1-Q3) | <br>14.9 ± 4.0<br>15 (12-18) | <br>16.1 ± 4.0<br>17 (13-19) | <br>17.5 ± 3.5<br>18 (16-20) | **p=0.0002**; **p=0.002***<br>**1 vs 2 p=0.03**<br>**1 vs 3 p=0.03**<br>**2 vs 3 p=0.01** |
| **BDI-II**<br>Mean ± sd<br>Median (Q1-Q3) | <br>10.8 ± 8.2<br>9 (5-14) | <br>14.2 ± 11.9<br>12.5 (5-19) | <br>13.9 ± 9.9<br>12 (6-20) | p=0.2; **p=0.01***<br>**1 vs 2 p=0.009**<br>**1 vs 3 p=0.04**<br>2 vs 3 p=0.6 |
| **EQ-5D-5L**<br>Mean ± sd<br>Median (Q1-Q3) | <br>69.2 ± 17.7<br>75 (62-80) | <br>72.6 ± 15.4<br>75 (67-85) | <br>66.7 ± 18.5<br>69 (53-80) | p=0.2; p=0.07* |

| | | | | |
|---|---|---|---|---|
| **ASR**<br>Total scale (Adaptative Functioning profile) N (%)<br>- Clinical range<br>- Limit range<br>- Normal range | <br><br>11 (16.7%)<br>15 (22.7%)<br>40 (60.6%) | <br><br>18 (35.3%)<br>5 (9.8%)<br>28 (54.9%) | <br><br>21 (38.2%)<br>7 (12.7%)<br>27 (49.1%) | **p=0.04**; p=0.1* |
| **Siegrist**<br>Effort-Reward imbalance, N (%) | 6 (8.6%) | 11 (18.0%) | 11 (16.4%) | p=0.2;p=0.7* |

**Supplementary table 3. Economic cost associated with days of work stoppage in patients with narcolepsy and controls**

This table present results in the 3 subgroups of patients according to age at disease onset and diagnosis, and sensitivity analyses considering (1) mean and median salary in the French population for all respondents rather than a salary based on socio-professional category (SPC) (2) patients with at least one sick leave day.
Note that loss of salary associated with full disability were not calculated given the low number of respondents on disability in both groups and the difficulty in obtaining a relevant valuation given the number of factors involved in the determination of the compensation amount.
Ref*: patients with NT1 onset and diagnosis < 18 years considered as the reference group

| | **Patients with NT1 (N=153)** | **Controls (N=124)** | **p-values adjusted for age and sex** | **Patients with NT1 with at least one sick leave day (N=40)** | **Controls with NT1 with at least one sick leave day (N=26)** | **p-values adjusted for age and sex** | **Patients with NT1 onset < 18 years** | | **Patients with NT1 onset ≥ 18 years (N=53)** (p-value adjusted for age and sex) |
|---|---|---|---|---|---|---|---|---|---|
| | | | | | | | **Diagnosis < 18 years (N=47)** (ref*) | **Diagnosis ≥ 18 years (N=53)** (p-value adjusted for age and sex) | |
| Number or sick leave days (mean ± sd) | 15.5 ± 51.4 | 8.7 ± 37.3 | p=0.096 | 59.3 ± 87.5 | 41.3 ± 74 | p=0.292 | 4.8 ± 20.0 (ref) | 7.6 ± 22.4 (p=0.725) | 33.0 ± 80.0 (p=0.283) |
| Annual loss of salary (income based on SPC) (mean ± sd €) | 736 ± 2922 | 473 ± 1788 | p<0.001 | 2798 ± 5210 | 2236 ± 3392 | p=0.517 | 188 ± 656 (ref) | 399 ± 1334 (p<0.001) | 1553 ± 4647 (p<0.001) |
| Annual loss of salary (income based on mean salary in French population) (mean ± sd €) | 622 ± 2005 | 377 ± 1515 | p=0.090 | 2363 ± 3368 | 1784 ± 2931 | p=0.365 | 216 ± 831 (ref) | 345 ± 962 (p=0.385) | 1253 ± 3085 (p=0.005) |
| Annual loss of salary (income based on median salary in French population) (mean ± sd €) | 427 ± 1349 | 260 ± 1019 | p=0.089 | 1623 ± 2249 | 1231 ± 1956 | p=0.360 | 150 ± 563 (ref) | 246 ± 664 (p=0.369) | 850 ± 2071 (p=0.006) |
